# Reliability of P3 Event-Related Potential during Working Memory Across the Spectrum of Cognitive Aging

**DOI:** 10.1101/2020.05.27.20109157

**Authors:** Hannes Devos, Jeffrey Burns, Ke Liao, Pedram Ahmadnezhad, Jonathan D. Mahnken, William M. Brooks, Kathleen Gustafson

## Abstract

Event-related potentials (ERPs) offer unparalleled temporal resolution in tracing distinct electrophysiological processes related to normal and pathological cognitive aging. The stability of ERPs in older individuals, who inherently show more intraindividual variability in cognitive functions, has not been established. In this test-retest reliability study, 39 older individuals (age 74.10 (5.4) years; 23 (59%) women; 15 non β-amyloid elevated, 16 β-amyloid elevated, 8 cognitively impaired) with scores on the Montreal Cognitive Assessment (MOCA) ranging between 3 and 30 completed a working memory (n-back) test with three levels of difficulty at baseline and two-week follow-up. Stability of the ERP was evaluated on grand averaged task effects for both visits. P3 peak amplitude and latency were measured in frontal channels. P3 peak amplitude at Fz, our main outcome variable, showed excellent reliability in 0-back (intraclass correlation coefficient (ICC), 95% confidence interval = 0.82 (0.67 – 0.90) and 1-back (ICC = 0.87 (0.76 – 0.93), however, only fair reliability in 2-back (ICC = 0.53 (0.09 – 0.75). Reliability of P3 peak latencies was substantially lower, with ICCs ranging between 0.17 for 2-back and 0.54 for 0-back. Generalized linear mixed models showed no confounding effect of age, group, or task difficulty on stability of P3 amplitude and latency of Fz. By contrast, MOCA scores tended to negatively correlate with P3 amplitude of Fz (p=0.07). We conclude that P3 peak amplitude and latency provide a stable measure of electrophysiological processes in older individuals. However, impaired cognition may affect the stability of the ERP response.

## 1 Introduction

The aging process is characterized by gradual decline in physical, neurobiological and cognitive functions that may impact instrumental activities of daily living (iADL) such as driving, doing household chores, managing finances, medication adherence, or grocery shopping.(1, 2) Deterioration in these iADL becomes more apparent with age-related neurodegeneration such as mild cognitive impairment (MCI) and Alzheimer’s disease (AD).(3) Executive functions in particular are paramount in carrying out numerous iADL, but are also vulnerable to the effects of normal and pathological cognitive aging.(4, 5) Working memory is one core executive function that relates to the ability to temporarily store, process, and manipulate the information necessary for higher order cognitive tasks such as decision making, learning, and reasoning.(6) Working memory stems from the interaction between attention, short-term retention and manipulation of information, carried out by the coordinated activation of many brain regions.(7)

The prefrontal cortex has particularly been associated with working memory.(8) Consequently, the prefrontal cortex is highly susceptible to the effects of aging and early neurodegeneration.(9, 10) A recent meta-analysis pooling functional magnetic resonance imaging studies suggested a gradual and linear decline in prefrontal cortex engagement in older individuals.(11) Similarly, electrophysiological processes also decline with age. The P3, a positive peak that appears with a latency between 250 to 500ms in the event-related potential (ERP), has been implicated in attention and working memory processes across the lifespan.(12) A previous study showed reduced positivity in P3 central-frontal and parietal ERPs in older adults,(13) whereas others demonstrated frontal hyperactivity in P3 coupled with parietal or posterior hypoactivity.(14, 15). Despite the ambiguity in ERP findings, most studies conclude that the abnormal ERP response in older individuals reflects inefficient or compensatory use of neural resources due to frontal cortex dysfunction.(13, 14) Therefore, electrophysiological responses to working memory tasks are convenient measures to test hypotheses related to frontal cortex function, normal cognitive aging, and early neurodegeneration.

The ability to distinguish natural variability and measurement error from biologically relevant cognitive changes due to aging or early neurodegeneration is valuable to provide informed decisions on diagnosis, monitoring, and treatment of cognitive impairments.(16) However, older adults show more intraindividual variability in performance measures of working memory compared to younger adults. The age-related changes in intraindividual variability of performance measures become even more apparent with increasing cognitive demand.(9) This increased intraindividual may also stem from the heterogeneity of cognitive profiles in older individuals, especially when patients with MCI and AD are included.(17) The intraindividual variability observed in performance measures is believed to be linked to frontal cortex dysfunction,(9) which may therefore also affect intraindividual variability of the ERP response in older adults.(18) To date, few studies have investigated test-retest reliability of P3 ERP in healthy older adults(19-21) The test-retest reliability of the P3 ERP in older individuals with a heterogeneous cognitive profile has yet to be established.

The aims were to (1) characterize test-retest reliability of P3 amplitude and latency, particularly in the frontal-midline channels; and (2) investigate the impact of age, disease groups, cognitive status, and task difficulty P3 amplitude and latency.

## 2 Materials and Methods

### 2.1 Participants

This test-retest reliability study included 39 right-handed participants recruited from the KU Alzheimer’s Disease Center between 05/03/2018 and 03/10/2020. Inclusion criteria were (1) age older than 65; (2) having undergone a previous amyloid PET scan of the brain; (3) ability to understand the instructions in English; and (4) informed consent. Exclusion criteria were: (1) currently taking steroids, benzodiazepines, or neuroleptics; (2) history of any substance abuse; and (3) history of a neurological disorder other than MCI or AD. Sixteen were cognitively normal older adults with no elevated amyloid PET scans (Aβ-), 15 were cognitive normal with elevated amyloid PET scans (Aβ+), and eight had a clinical diagnosis of mild cognitive impairment with positive amyloid PET scans. Participants completed their two-week follow-up session 16 ± 8 days after the first session. Each session lasted about 60 minutes including rest breaks.

### 2.2 Procedure

#### 2.2.1 Demographic and Clinical Information

Age, sex, and education were recorded. General cognitive functions were evaluated with the Montreal Cognitive Assessment (MOCA).(22) Scores on the MOCA range between 0 and 30.

#### 2.2.2 N-back Test

In the n-back test, participants are shown a series of letters and are instructed to press a button when the current stimulus is the same as the item presented n-positions back. The cognitive demand of the n-back task increases with each number, while the perceptual and motor demands remain constant. Whereas in the 0-back and 1-back the stimulus on screen is held in the focus of attention, the more difficult versions of the n-back tests require constant switching from the focus of attention to short- term memory.(23) Higher levels of difficulty require continuous mental effort to update information of new stimuli and maintain representations of recently presented stimuli.(24)

In this study, the 0-back, 1-back, and 2-back tests were administered. Participants sat in a comfortable chair at 26 inches in front of the computer screen with the center of the screen at eye level. White letters appeared on a black screen. Prior to each test, participants were given a practice trial consisting of 7 non-targets and 3 targets. The practice trials were repeated until the participant felt comfortable with the instructions. Each test comprised 180 trials, including 60 trials that needed a response (target, 33.3%) and 120 trials for which a response was not required (nontarget, 66.7%). Each letter was presented for 500 ms on the computer screen followed by a blank interstimulus interval for 1700 ms, with a random jitter of +/-50 ms. The maximum time to accept the response was 2150 ms. The total task time was ∼7 minutes. In the 0-back test, participants were instructed to press the left mouse button as soon as the letter “X” (target) appeared on the screen while ignoring the other letters (nontarget). In the 1-back test, participants were instructed to press the button if the current letter on the screen was the same as the letter previously shown (target). In the 2-back test, participants were instructed to press the button when the current letter was the same as the one presented two places before (target). The number of hits (accuracy) and response times to the hits were the main behavioral performance outcome measures.

#### 2.2.3 P3 ERP

Continuous electro-encephalogram (EEG) was acquired using a Philips EGI high-density system from 256 scalp electrodes, digitized at 1,000 Hz. Data were filtered from 0.50 to 30 Hz using EGI software. Data were online referenced to Cz and offline rereferenced to the left and right mastoid. All other EEG processing was done in EEGLab(25) and in ERPLab.(26) Various artifacts unrelated to cognitive functions, including ocular and muscular movement, cardiovascular signal were identified and removed using independent component analysis (ICA). Signal from bad electrodes were interpolated using surrounding electrode data. Stimulus-locked ERPs were extracted from the n-back tests and segmented into epochs of 100 ms before to 1000ms after stimulus onset, and baseline corrected using the prestimulus interval. Scalp locations and measurement windows for the P3 component were based on their spatial extent and latency after inspection of grand average waveforms (collapsed across the two sessions). P3 peak amplitude and peak latency of the task effect were considered the main electrophysiological outcome measures. The difference waveform was calculated by subtracting the average ERP elicited from the targets from the average ERP elicited by non-targets for each participant. The P3 component time window was established between 200 ms and 400 ms for all three tests. Because of the prefrontal cortex involvement in working memory, we identified a priori the Fz channel as the main channel, but also calculated reliability of other pre-identified electrode locations, i.e., Cz, Pz, F3, and F4. Cz was interpolated using the surrounding five channels. No participants were removed from the analyses because of artifacts. However, one participant disengaged during the 2-back test and was therefore excluded from the analyses.

### 2.3 Data Analysis

Descriptive analysis including mean (standard deviation) and frequency count of participants’ general, performance measures, and ERP data were performed as appropriate. Intra-class correlation coefficients (ICC) were used to calculate test-retest reliability of performance measures and P3 amplitude and latency. ICCs reflect the consistency of a measure taking into account variance related to the time of testing.(27) ICC values less than 0.40 were considered poor; values between 0.40 and 0.59 fair, values between 0.60 and 0.74 good, and values between 0.75 and 1.00 excellent.(28). Bland-Altman plots were used to visualize the measurement precision of amplitude and latency across the test moments.(29) Intersubject stability according to subject rankings was calculated using the Pearson r correlation coefficient. Generalized linear mixed models were employed to evaluate the effect of age, diagnosis (Aβ-; Aβ+; MCI/AD), MOCA scores, and task difficulty on stability of the P3 amplitude and latency. Stability of P3 amplitude (latency) was calculated as the squared difference of P3 amplitude (latency) at follow-up and baseline. The Kolmogorov–Smirnov test was employed to test the normality of our data distribution in addition to visualization of Q-Q plots. All analyses were done using SAS 9.4 software. The threshold of significance was set at p = 0.05.

## 3 Results

### 3.1 Participant Characteristics

Participants (n = 39) were on average 74.05 (5.37) years old and scored 26.44 (4.76) on the MOCA scale. However, MOCA scores ranged from 3 to 30, indicating a wide range of severe to normal cognitive functions. About 60% (n = 23) were women.

### 3.2 Test-Retest Reliability of Performance Measures

All ICC values of hits (accuracy) and response times of each n-back test demonstrated excellent reliability (Supplementary Table 1). ICCs of hits ranged between 0.92 (1- and 2-back) and 0.99 (0- back) and were slightly higher than the ICCs of response times, ranging between 0.76 (2-back) and 0.89 (1-back). Pearson r correlations ranged from 0.65 (0-back response time) to 0.99 (0-back hits).

### 3.3 Test-Retest Reliability of ERP Measures

Grand average waveforms of the task effect from all channels at baseline and follow-up are displayed in Figure 1. The 3D scalp map is embedded in the figure to demonstrate the task effect at P3. Considerable overlap in ERP response within the P3 time window (200 – 400 ms post-stimulus) was observed at baseline and two-week follow-up.

**Figure 1.**
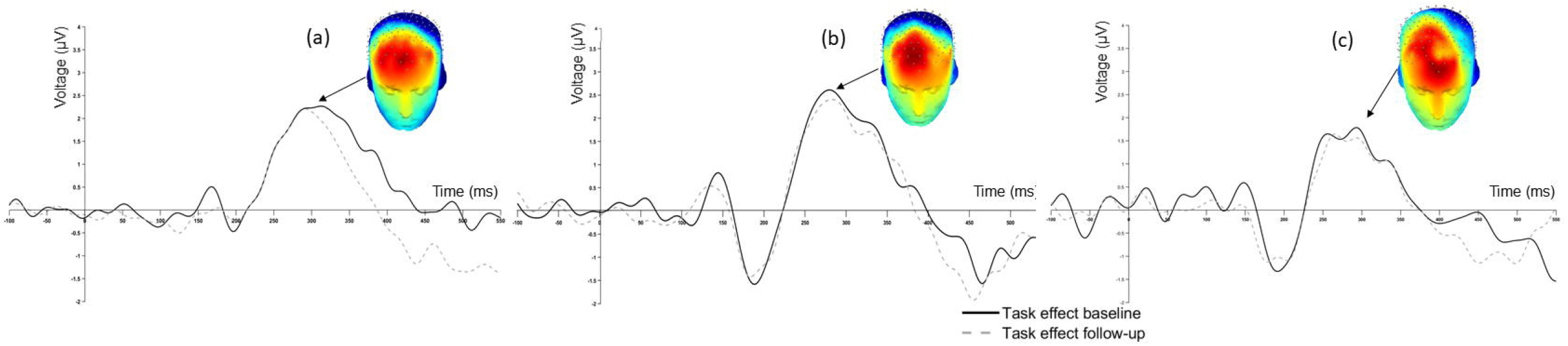
Grand Average Event-related Potential Waveform of (a) 0-back, (b) 1-back and (c) 2-back.

The ICC values of P3 peak amplitude and peak latency of the key electrode locations are displayed in Table 1. Overall, P3 amplitude showed greater reliability compared to P3 latency across channels and task difficulty levels. Also, ICCs of the 0-back and 1-back were consistently higher than those calculated for the 2-back.

**Table 1.**
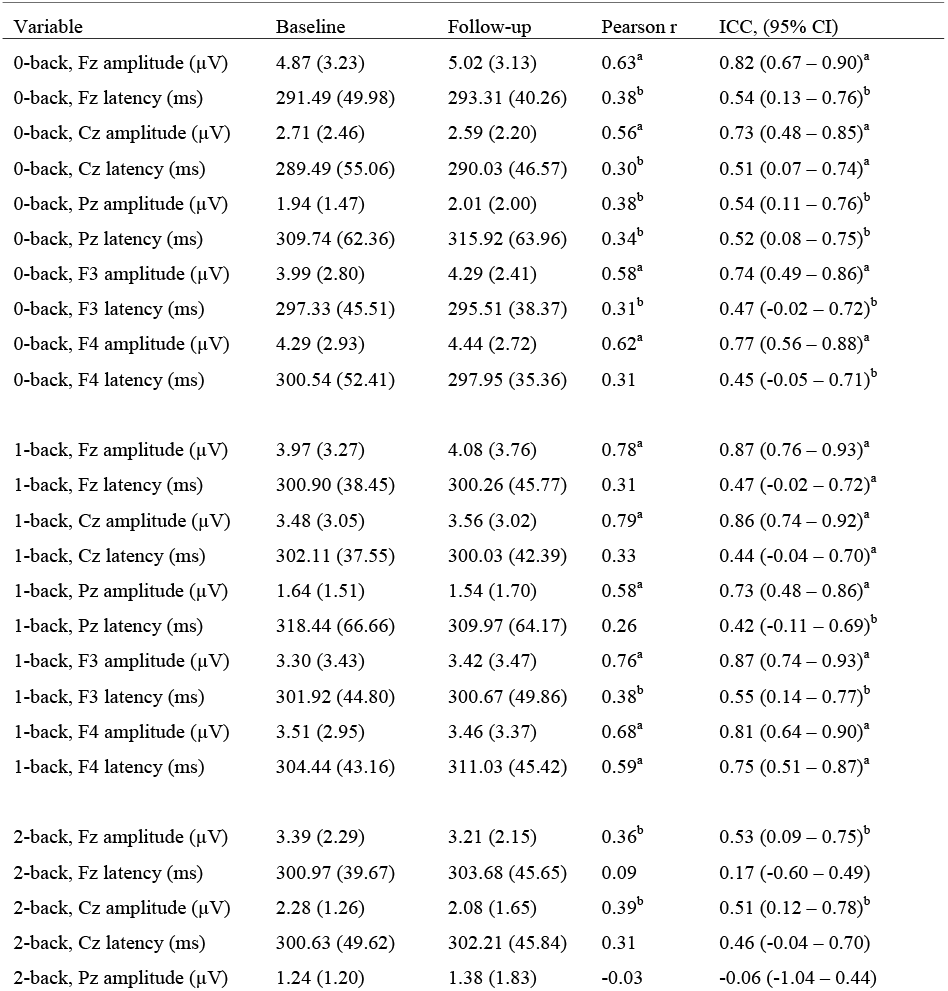
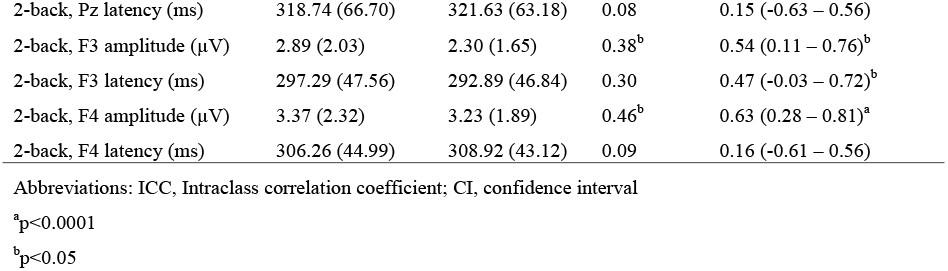
Comparison of task effect (target – nontarget) P3 response at baseline and two-week follow-up.

| Variable | Baseline | Follow-up | Pearson r | ICC, (95% CI) |
| --- | --- | --- | --- | --- |
| 0-back, Fz amplitude (μV) | 4.87 (3.23) | 5.02 (3.13) | 0.63<sup>a</sup> | 0.82 (0.67 – 0.90)<sup>a</sup> |
| 0-back, Fz latency (ms) | 291.49 (49.98) | 293.31 (40.26) | 0.38<sup>b</sup> | 0.54 (0.13 – 0.76)<sup>b</sup> |
| 0-back, Cz amplitude (μV) | 2.71 (2.46) | 2.59 (2.20) | 0.56<sup>a</sup> | 0.73 (0.48 – 0.85)<sup>a</sup> |
| 0-back, Cz latency (ms) | 289.49 (55.06) | 290.03 (46.57) | 0.30<sup>b</sup> | 0.51 (0.07 – 0.74)<sup>a</sup> |
| 0-back, Pz amplitude (μV) | 1.94 (1.47) | 2.01 (2.00) | 0.38<sup>b</sup> | 0.54 (0.11 – 0.76)<sup>b</sup> |
| 0-back, Pz latency (ms) | 309.74 (62.36) | 315.92 (63.96) | 0.34<sup>b</sup> | 0.52 (0.08 – 0.75)<sup>b</sup> |
| 0-back, F3 amplitude (μV) | 3.99 (2.80) | 4.29 (2.41) | 0.58<sup>a</sup> | 0.74 (0.49 – 0.86)<sup>a</sup> |
| 0-back, F3 latency (ms) | 297.33 (45.51) | 295.51 (38.37) | 0.31<sup>b</sup> | 0.47 (-0.02 – 0.72)<sup>b</sup> |
| 0-back, F4 amplitude (μV) | 4.29 (2.93) | 4.44 (2.72) | 0.62<sup>a</sup> | 0.77 (0.56 – 0.88)<sup>a</sup> |
| 0-back, F4 latency (ms) | 300.54 (52.41) | 297.95 (35.36) | 0.31 | 0.45 (-0.05 – 0.71)<sup>b</sup> |
| 1-back, Fz amplitude (μV) | 3.97 (3.27) | 4.08 (3.76) | 0.78<sup>a</sup> | 0.87 (0.76 – 0.93)<sup>a</sup> |
| 1-back, Fz latency (ms) | 300.90 (38.45) | 300.26 (45.77) | 0.31 | 0.47 (-0.02 – 0.72)<sup>a</sup> |
| 1-back, Cz amplitude (μV) | 3.48 (3.05) | 3.56 (3.02) | 0.79<sup>a</sup> | 0.86 (0.74 – 0.92)<sup>a</sup> |
| 1-back, Cz latency (ms) | 302.11 (37.55) | 300.03 (42.39) | 0.33 | 0.44 (-0.04 – 0.70)<sup>a</sup> |
| 1-back, Pz amplitude (μV) | 1.64 (1.51) | 1.54 (1.70) | 0.58<sup>a</sup> | 0.73 (0.48 – 0.86)<sup>a</sup> |
| 1-back, Pz latency (ms) | 318.44 (66.66) | 309.97 (64.17) | 0.26 | 0.42 (-0.11 – 0.69)<sup>b</sup> |
| 1-back, F3 amplitude (μV) | 3.30 (3.43) | 3.42 (3.47) | 0.76<sup>a</sup> | 0.87 (0.74 – 0.93)<sup>a</sup> |
| 1-back, F3 latency (ms) | 301.92 (44.80) | 300.67 (49.86) | 0.38<sup>b</sup> | 0.55 (0.14 – 0.77)<sup>b</sup> |
| 1-back, F4 amplitude (μV) | 3.51 (2.95) | 3.46 (3.37) | 0.68<sup>a</sup> | 0.81 (0.64 – 0.90)<sup>a</sup> |
| 1-back, F4 latency (ms) | 304.44 (43.16) | 311.03 (45.42) | 0.59<sup>a</sup> | 0.75 (0.51 – 0.87)<sup>a</sup> |
| 2-back, Fz amplitude (μV) | 3.39 (2.29) | 3.21 (2.15) | 0.36<sup>b</sup> | 0.53 (0.09 – 0.75)<sup>b</sup> |
| 2-back, Fz latency (ms) | 300.97 (39.67) | 303.68 (45.65) | 0.09 | 0.17 (-0.60 – 0.49) |
| 2-back, Cz amplitude (μV) | 2.28 (1.26) | 2.08 (1.65) | 0.39<sup>b</sup> | 0.51 (0.12 – 0.78)<sup>b</sup> |
| 2-back, Cz latency (ms) | 300.63 (49.62) | 302.21 (45.84) | 0.31 | 0.46 (-0.04 – 0.70) |
| 2-back, Pz amplitude (μV) | 1.24 (1.20) | 1.38 (1.83) | -0.03 | -0.06 (-1.04 – 0.44) |
| Test-Retest Reliability of ERP in Older Adults |  |  |  |  |
| 2-back, Pz latency (ms) | 318.74 (66.70) | 321.63 (63.18) | 0.08 | 0.15 (-0.63 – 0.56) |
| 2-back, F3 amplitude (μV) | 2.89 (2.03) | 2.30 (1.65) | 0.38<sup>b</sup> | 0.54 (0.11 – 0.76)<sup>b</sup> |
| 2-back, F3 latency (ms) | 297.29 (47.56) | 292.89 (46.84) | 0.30 | 0.47 (-0.03 – 0.72)<sup>b</sup> |
| 2-back, F4 amplitude (μV) | 3.37 (2.32) | 3.23 (1.89) | 0.46<sup>b</sup> | 0.63 (0.28 – 0.81)<sup>a</sup> |
| 2-back, F4 latency (ms) | 306.26 (44.99) | 308.92 (43.12) | 0.09 | 0.16 (-0.61 – 0.56) |

Abbreviations: ICC, Intraclass correlation coefficient; CI, confidence interval
<sup>a</sup>p<0.0001
<sup>b</sup>p<0.05

For the main channel location Fz, excellent reliability was found in P3 amplitude for 0-back (ICC = 0.82) and 1-back (ICC = 0.87). P3 amplitude of Fz for 2-back only showed fair reliability. Reliability scores of P3 latency at Fz were fair for 0-back (ICC = 0.54) and 1-back (ICC = 0.47), but poor for 2- back (ICC = 0.17). Figure 2 shows the Bland-Altman plots for P3 peak amplitude and peak latency at the Fz channel. All plots demonstrated equal distribution of the data around zero, indicating no bias in the results and no heteroscedasticity within the data.

**Figure 2.**
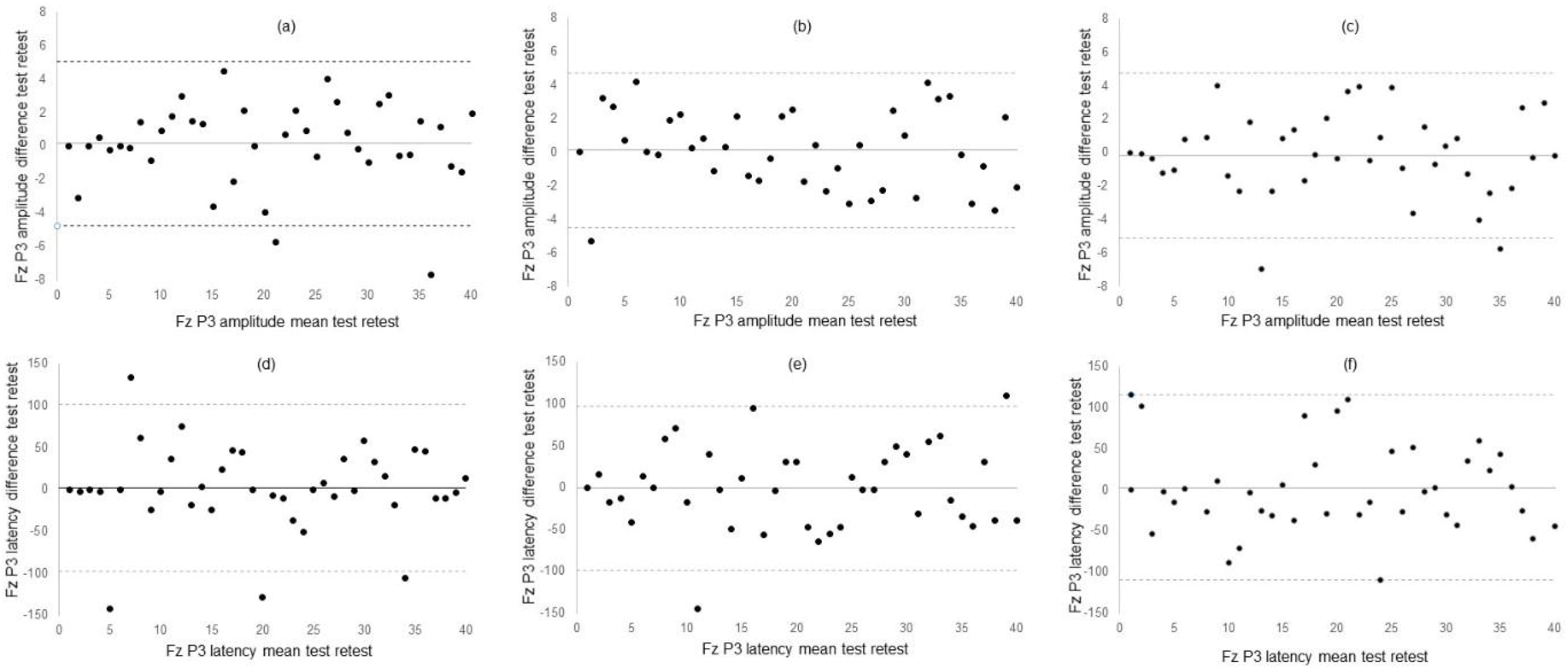
Bland Altman Plotsof (a) 0-back Fz Peak Amplitude (b) 1-back Fz Peak Amplitude; (c) 2- back Fz Peak Amplitude; (d) 0-back Fz Peak Latency; (e) 1-back Peak Latency; (f) 2-back Peak Latency.

Finally, generalized linear mixed models were employed to evaluate the effect of age, disease diagnosis (Aβ-; Aβ+; MCI/AD), cognitive status, and task difficulty on stability of squared P3 peak amplitude and latency at the Fz channel. Age (p=0.74), disease diagnosis (p = 0.67), and task difficulty (p = 0.70) did not affect the stability of the P3 amplitude response, although individuals with lower MOCA cognitive scores tended to show more variability in P3 amplitude (p = 0.07). Age (p = 0.60), disease diagnosis (p = 0.55), MOCA (p = 0.52), or task difficulty (p = 0.95) did not affect the stability of the P3 latency response.

We recalculated ICC for 0-back, 1-back, and 2-back in participants who scored 26 or higher on MOCA (n = 32) and those scoring lower than 26 (n = 7). ICC values showed more variance in 0-back and in 2-back in the group with lower MOCA scores, but ICC values were not worse across the n-back tests in this group (Supplementary Tables 2 and 3). Whereas ICCs were similar in the Aβ- and Aβ+ groups, lower ICCs were found for the MCI/AD group (Supplementary Tables 2 and 3).

## 4 Discussion

This test-retest reliability study provides critical information on the stability of electrophysiological measures related to working memory in healthy older adults, older adults with increased risk of dementia, and those with MCI or AD. Our results showed that most P3 ERPs in the frontal channels provide fair to excellent reliability to measure electrophysiological processes of cognitive aging in older adults with and without cognitive impairments. Similar to previous studies, the reliability is superior in measures of amplitude compared to latency.(20, 21, 30, 31) The robustness of P3 stability is not affected by age, disease diagnosis, or task difficulty, however, there is a trend that lower MOCA scores may affect the stability of the P3 amplitude response.

The body of evidence related to reliability of P3 ERPs is sparse, and typically restricted to healthy young,(30-34) middle-aged,(31) and older individuals.(19-21). Few studies have reported reliability measures in neurological conditions.(35) The reliability analyses in our study produced fair to excellent ICC values across the n-back tests. Whereas ICC values provide a single measure of the magnitude of agreement, Bland-Altman plots depict a graphical display of bias across the two test moments.(36) Visual inspection of the Bland-Altman plots showed average difference in ERP responses between first and second testing close to 0, with equal spread of data points around the average difference line. These findings suggest that 2 weeks follow-up is sufficient to wash out any potential adaptation, test, or practice effect of the n-back on ERPs in older individuals.

Comparison of our results with other test-retest studies of ERPs in older adults is complicated by lack of consistency in terms of the ERP components that are investigated, the tests of working memory, the choice of channel locations, the extracted P3 metric, the P3 window measurement, and the test- retest reliability intervals.(19-21). Our research design most closely aligns with a study that compared ERPs to novel stimuli collected at baseline and 7-week follow-up in healthy older individuals.(21) Similar to our study, this study also found excellent reliability for P3 mean amplitude (ICC =0.86, 95% CI, 0.78 – 0.92), and poorer reliability for P3 mean latency (ICC = 0.56, 0.30 – 0.73). Our study demonstrated larger confidence intervals in some of the amplitude and latency measures, which might have been due to the greater cognitive heterogeneity of our sample. Another reliability study also reported considerably lower reliability in P3 amplitude (ICC = -0.02) and latency (ICC = -0.17) in seven individuals experiencing cognitive difficulties following traumatic brain injury compared to healthy peers (ICC = 0.84 for amplitude and 0.64 for latency).(35) Combined, these findings point towards a potential confounding effect of cognitive impairment on stability of ERPs in neurological conditions.

No effect of age, task difficulty, or disease diagnosis was found on stability of the P3 ERP in the n- back task. Most participants in our study were cognitively normal, either without (n = 15) or with (n = 16) elevated beta amyloid. Accumulation of beta amyloid deposits in the brain is known to increase the risk of developing AD.(37) The fair to excellent reliability of P3 amplitude and latency in our group of older adults provides opportunities for studying the effect of amyloid deposits on neural transmission in preclinical AD using ERP. P3 amplitudes are smaller in AD compared to controls.(38) ERPs also show useful in predicting conversion to AD, with accuracy rates ranging between 70% and 94%.(39) Patients with AD exhibit prolonged latency in P3 ERP compared to age-matched controls(40). These prolonged latencies observed in patients with AD become particularly apparent in the cognitive domains of executive function, memory, and language.(41) The ability of P3 ERP to discriminate between mild cognitive impairment (MCI) and AD(42) opens avenues for investigation of ERP in detecting preclinical AD.(43)

Although this study included participants with a wide range of cognitive impairment, most were cognitively normal. Future studies should include a larger sample of participants with MCI and AD to confirm the confounding effect of impaired cognition on the stability of the P3 response. Although the n-back is arguably the most ubiquitous working memory test used in ERP studies across the age spectrum,(23) we did not establish reliability of ERP in other cognitive domains known to deteriorate in older age, such as memory and language, and this remains an opportunity for further investigation. Our results are therefore only generalizable to the specific subdomain of working memory. Future research should also include multiple testing sessions over extended periods of time to evaluate the sensitivity of ERP to detect subtle neurobiological changes due to normal and pathological aging.

## Data Availability

The data are available upon request.

## 5 Conclusion

We set out to assess the test-retest reliability of ERP response in older adults with a heterogeneous cognitive profile. Consistent with other studies, P3 amplitude and latency show fair to excellent reliability across different levels of task difficulty. However, impaired cognition may potentially affect the stability of the P3 ERP response.

## Ethics Statement

The study was carried out in accordance with the Declaration of Helsinki. The study was approved by the University of Kansas Medical Center Internal Review Board. Informed consent was obtained from each participant. Each participant received $100 for participating.

## Author Contributions

HD, JB, JM, WMB, and KG conceptualized the study. HD, KL, and KG worked out the EEG data processing steps. HD, PA, and KL administered the tests. HD and JM analysed the data. HD wrote the initial manuscript. JB, KL, PA, JM, WMB, and KG reviewed the manuscript and provided valuable comments.

## Acknowledgments

The authors thank the volunteers for their time and willingness to participate in this research. We are also grateful for the staff at the KU Alzheimer Disease Center.

